# Quantum-Enhanced Transfer Learning for MS Lesion Detection in MRI: A Hybrid Classical-Quantum Framework

**DOI:** 10.1101/2025.11.16.25340339

**Authors:** Mohadeseh Zarei Ghobadi, Elaheh Afsaneh

## Abstract

Accurate detection of multiple sclerosis (MS) lesions in magnetic resonance imaging (MRI) requires robust deep learning models to capture subtle spatial and textural features. We introduce hybrid quantum-classical transfer learning algorithms for MS classification using axial and sagittal MRI scans, combining classical convolutional neural networks (CNNs) including EfficientNetB3, ResNet50, DenseNet121 with parameterized quantum circuits to enhance feature representation via entanglement and quantum-specific non-linearities. Quantum layers are trained end-to-end with classical backbones via backpropagation, enabling seamless integration of quantum-enhanced features. For axial MRI, QResNet50 achieved a high accuracy of 97.58% and AUC of 99.31%, while QDenseNet121 reached 97.28% accuracy and 99.13% AUC. For sagittal MRI, classical ResNet50 excelled with 99.15% accuracy and 99.93% AUC, while QEfficientNetB3 improved accuracy (97.46% to 98.30%) but reduced AUC (99.51% to 99.32%), and QDenseNet121 achieved 98.87% accuracy and 99.83% AUC. Hybrid models showed mixed results, with QCNN underperforming, suggesting quantum benefits are architecture-dependent. Despite simulated quantum circuits mitigating hardware limitations, our results demonstrate the potential to enhance diagnostic performance in specific architectures. This work clarifies a foundational step toward quantum-enhanced deep learning for clinical applications, opening research directions in quantum-aware transfer learning and error mitigation for biomedical imaging.

## 1 Introduction

Quantum machine learning (QML), which lies at the intersection of quantum computing and classical machine learning, has undergone significant advancements in recent years due to the unique strengths of quantum technology in enhancing data processing and model training (1). However, current hardware limitations, such as limited qubit counts and error rate, continue to pose challenges for the full realization and scalability of quantum models (2, 3). Quantum neural networks (QNNs), a key component of QML, have demonstrated promising results across various domains by utilizing quantum phenomena like superposition and entanglement to improve learning performance. Among the various quantum neural network architectures, Quantum Convolutional Neural Networks (QCNNs) have emerged as a powerful hybrid quantum-classical approaches (4). QCNNs utilize the potential of quantum computing inspired by classical convolutional neural networks to reduce dimensionality and tackle complex high-dimensional problems. This approach is especially valuable in healthcare applications and medical diagnostics, particularly for the early detection of Multiple Sclerosis (MS), where timely and accurate diagnosis is critical.

Multiple Sclerosis is a chronic inflammatory disorder of the central nervous system characterized by damage to the myelin sheath and nerve fibers (5). The myelin sheath is a fatty-protein coating that insulates nerve cells in the brain and spinal cord, enabling rapid transmission of electrical impulses (6). Damage to this protective layer disrupts nerve signal conduction and leads to the formation of scar tissue, known as plaques or sclerosis.

The number of people suffering from this disease is estimated to be approximately 2-3 million worldwide (7). Multiple sclerosis is more commonly observed in young to middle-aged females, although this has not always been the case (8). The etiology of MS remains unclear, but it is widely recognized as a multifactorial disease influenced by genetic and environmental risk factors (9, 10). The pathology of MS is highly heterogeneous, posing challenges in accurately diagnosing the disease in its early stages, measuring progression, and evaluating treatment efficacy (8). Its diagnosis remains difficult due to the absence of specific symptoms, physical findings, or laboratory tests that can definitively confirm the condition (11). Consequently, physicians rely on a combination of diagnostic methods, including thorough review of the patient’s medical history, neurological examinations, magnetic resonance imaging (MRI), cerebrospinal fluid analysis, and blood tests (10, 12–15) to exclude other conditions that present with similar symptoms. Among these, MRI modalities are considered the most effective non-invasive tool for detecting MS lesions, providing crucial information about brain structure and tissue abnormalities. MRI biomarker, such as lesion count, evolution over time, and brain volume measurements (including grey matter and white matter) are valuable indicators for disease prognosis and treatment response (16–19). To enhance diagnostic accuracy, boost efficiency, and minimize manual errors, artificial intelligence (AI)-powered computer-aided diagnosis systems have been integrated with conventional MRI technology.

Recent advancements in AI, particularly deep learning (DL) methods like convolutional neural networks (CNNs), has significantly enhanced the accuracy and efficiency of diagnosing MS through MRI data analysis (20, 21). In a study, multiple sclerosis patients were categorized based on their disability level using a deep learning model incorporating convolutional neural networks (22). The model outperformed traditional methods by effectively extracting optimal MRI features. Attention-map analysis showed the frontotemporal cortex and cerebellum play crucial roles in predicting disability and also indicated that MS progression involves a complex distribution of neural damage across the central nervous system. In another study, a four-layer deep belief network was used to extract latent features from normal-appearing white and gray matter image patches, selected via voxel-wise t-tests and excluding lesion-overlapping areas (23). These features were then undergone LASSO-based selection to reduce overfitting and are used to train a random forest classifier to distinguish multiple sclerosis patients from healthy controls. The integration of multimodal myelin and T1-weighted features enhanced classification accuracy compared to single-modality approaches, highlighting the advantage of joint feature learning in improving predictive performance.

To improve the accuracy of multiple sclerosis diagnosis using MRI scans, a model was developed that integrates a multi-view ResNet architecture with novel attention mechanisms-the View Space Attention Block (VSAB) and View Channel Attention Block (VCAB)-to extract detailed features from 2D brain images. Additionally, the Quantum RIME (QRIME) algorithm was introduced, combining RIME with Quantum Behaved Particle Swarm Optimization (QPSO) to perform efficient dimensionality reduction, thereby enhancing both diagnostic accuracy and computational efficiency (24). The results showed the potential of AI applications in improving the early detection of neurodegenerative diseases. In another study, disability progression prediction in MS patients was investigated using a time-dependent deep learning model based on sequential medical imaging data (25). The study captured temporal relationships across multiple MRI timepoints and demonstrated that quantum models achieved competitive performance compared to classical neural network architectures in binary classification of MS disability.

In this study. we explored the application of classical and hybrid quantum-classical deep learning models for the detection of MS lesions in axial and sagittal brain MRI scans. Given the heterogeneous spatial distribution and textural characteristics of MS lesions, the ability of these models to adapt to such variability plays a crucial role in achieving reliable and robust diagnostic performance. Leveraging transfer learning techniques with architectures such as ResNet50, EfficientNetB3, and DenseNet121, we established a classical baseline before integrating parameterized quantum circuits to form hybrid quantum-classical models. A comparative evaluation across imaging planes revealed distinct performance trends, showing both the promise and current limitations of quantum-enhanced deep learning in the context of medical image analysis.

This study is systematically organized to investigate hybrid quantum-classical approaches for MS detection. The Introduction highlights the clinical importance of MS diagnosis and reviews the application of deep learning methods in improving diagnostic accuracy using MRI data. The Methods section provides a rigorous technical exposition of: (i) classical CNN architectures (CNN, DenseNet121, ResNet50, EfficientNetB3) and their quantum hybrid variants, (ii) quantum circuit design principles including amplitude/angle embedding strategies, and (iii) the experimental framework encompassing data preprocessing and hyperparameter optimization. The Results section presents a comprehensive performance analysis across axial and sagittal MRI planes, with quantitative evaluation through accuracy, precision, recall, F1-score, and AUC metrics. The Discussion explains these findings within current literature, examines the differences between quantum and classical performance, and addresses practical implementation constraints. Finally, the Conclusion synthesizes key insights regarding quantum-enhanced feature learning while outlining future research directions in quantum medical imaging.

## 2 Theoretical Background

### 2.1 Quantum Computing

In this section, we provide a brief background and introduce fundamental quantum computing concepts. These preliminaries will be used to describe the QCNN algorithms in the following sections.

#### Quantum bit (qubit)

A qubit is mathematically described as a vector in a two-dimensional Hilbert space, with the computational basis states |0⟩ and |1⟩ forming its foundation (26). Unlike classical bits, qubits can exist in a superposition state, meaning they can simultaneously represent a linear combination of these basis states weighted by complex probability amplitudes. This property enables quantum entanglement, where the states of several qubits become connected so that the state of each qubit depends on the others, no matter how far apart they are. When one qubit in an entangled system is measured, its state collapses and immediately determines the states of the others. These extraordinary features allow entangled qubits to encode and process information in ways that far exceed the capabilities of classical bits, giving quantum computers the potential to solve certain complex problems much more efficiently than traditional computers.

#### Quantum gates

Quantum gates are mathematical operations that act on one or more qubits to change their quantum states (27). The most commonly used gates include the Hadamard, CNOT, Rotation, and Pauli gates. The Hadamard gate creates superposition by transforming a qubit from a definite state into a combination of |0⟩ and |1⟩ states, while the CNOT gate as a two-qubit gate generates entanglement between qubits. Rotation gates perform parameterized rotations around the *X*. *Y*. or *Z* axes of the Bloch sphere, and Pauli gates (*X*. *Y*. and *Z*) are used for controlled bit and phase flips.

#### Quantum circuits

Quantum circuits consist of sequences of quantum gates arranged according to quantum algorithms to manipulate qubits (28). These circuits generate superposition and entanglement, which serve as fundamental resources for quantum computation. Quantum gates and circuits form the foundational elements in designing quantum computing models. Within a quantum circuit, measurement gates convert quantum states into classical information by collapsing them. Typically, several measurements are conducted, and their results are aggregated to estimate observable values and make predictions (29).

### 2.2 Quantum neural network (QNN)

A quantum neural network in contrast to a classical convolutional neural network, consists of quantum layers that typically include three main components: embedding (data encoding), quantum circuits (often variational circuits with parameterized gates and entanglement), and measurement (30). Initially, classical data is encoded into a quantum state via a state preparation routine or feature map, which is usually designed to enhance model performance (31). Following encoding, a variational quantum circuit with parameterized gates is applied and optimized for a specific task through loss function minimization (32). The output of the quantum model is then obtained by measuring the quantum state and applying classical post-processing to the measurement outcomes. This framework allows QNNs to leverage quantum phenomena to potentially improve computational efficiency and representational capacity compared to classical neural networks.

### 2.3 Quantum Encoding

Quantum machine learning relies on embedding classical data into quantum states using quantum principles (33). This process involves mapping data vectors into parameters that define quantum circuits, thereby generating quantum feature representations in Hilbert space. Two common encoding methods are amplitude encoding and angle encoding.

#### Amplitude encoding

In amplitude encoding, classical data is embedded into the probability amplitudes of a quantum state. By exploiting quantum superposition, the encoded information is accessed by measuring the quantum state’s amplitudes (34). A key advantage is its efficiency in representing large datasets using relatively few qubits. Specifically, encoding a classical dataset with *M* data points and *N* features per point requires *n* = *log*_2_(*NM*) qubits.

To perform amplitude encoding, classical data vectors *x* = (*x*_1_. *x*_2_. …. *x*_*N*_) are mapped to quantum states as:

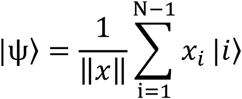

where |*i*⟩ denotes the computational basis states and *x*_*i*_ represents the *i*-th component of the classical data. The quantum system is initially prepared in the all-zero state |0⟩^⊗*n*^. where *n* is the number of qubits determined by the dataset size. A sequence of unitary operations, such as rotations and controlled gates, is then applied to transform this initial state into the desired encoded quantum state.

#### Angle encoding

Angle encoding embeds classical data into quantum states via parameterized rotations around the *X*. *Y*. or *Z* axes, with angles corresponding to data values. For example, a classical data vector *x* = (*x*_1_. *x*_2_. …. *x*_*N*_) can be encoded using rotations around the *Y*-axis as:

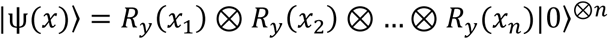

Angle encoding maps each feature to the rotation angle of a single qubit, requiring one qubit per feature. This method uses relatively fewer quantum gates, enhancing its compatibility with existing quantum hardware. Generally, encoding a dataset with n features requires exactly n qubits.

For datasets with a large number of features, amplitude encoding requires fewer qubits per data point but demands a higher number of quantum gates to prepare the corresponding quantum state. In contrast, angle encoding is simpler and more resilient to noise on current quantum devices, making it more practical given existing hardware limitations. However, angle encoding is less efficient when handling high-dimensional data. Each method offers distinct advantages and trade-offs, where the choice between amplitude and angle encoding depends on the available quantum hardware capabilities and the specific requirements of the machine learning task (35).

### 2.4 Circuit Design

The circuit design defines the sequence and types of quantum gates applied to the qubits after data encoding (36). Typically, this involves a combination of parameterized rotation gates and entangling gates applied to neighboring qubits. Entangling gates create quantum correlations across adjacent qubits, enabling the circuit to exploit quantum phenomena such as superposition and interference. This enhanced representational power enables quantum convolutional layers to capture more complex features from imaging data. During training, the parameters of the rotation gates are optimized to minimize a loss function, tailoring the quantum circuit to the specific learning task.

### 2.5 Measurement and Output Features

After processing through the quantum convolutional layer, measurements are performed on the qubits to extract expectation values of specific observables. These quantum-derived features constitute the output of the quantum layer and can be fed into subsequent layers-either quantum or classical-enabling hybrid quantum-classical architectures.

## 3 Methods

### 3.1 Dataset details and preprocessing

We utilized a dataset comprising 1441 axial and 2016 sagittal FLAIR MRI brain images exhibiting visible MS lesions, alongside 2016 axial and sagittal image slices depicting normal brain appearance without white matter lesions (37). The images prospectively collected from 72 patients diagnosed with MS and 59 healthy control subjects. Fig. 1 presents ten representative samples, with randomly selected axial images from the MS and healthy groups shown in Fig. 1a and Fig. 1b, respectively, and sagittal images from the MS and healthy groups displayed in Fig. 1c and Fig. 1d, respectively.

**Fig. 1.**
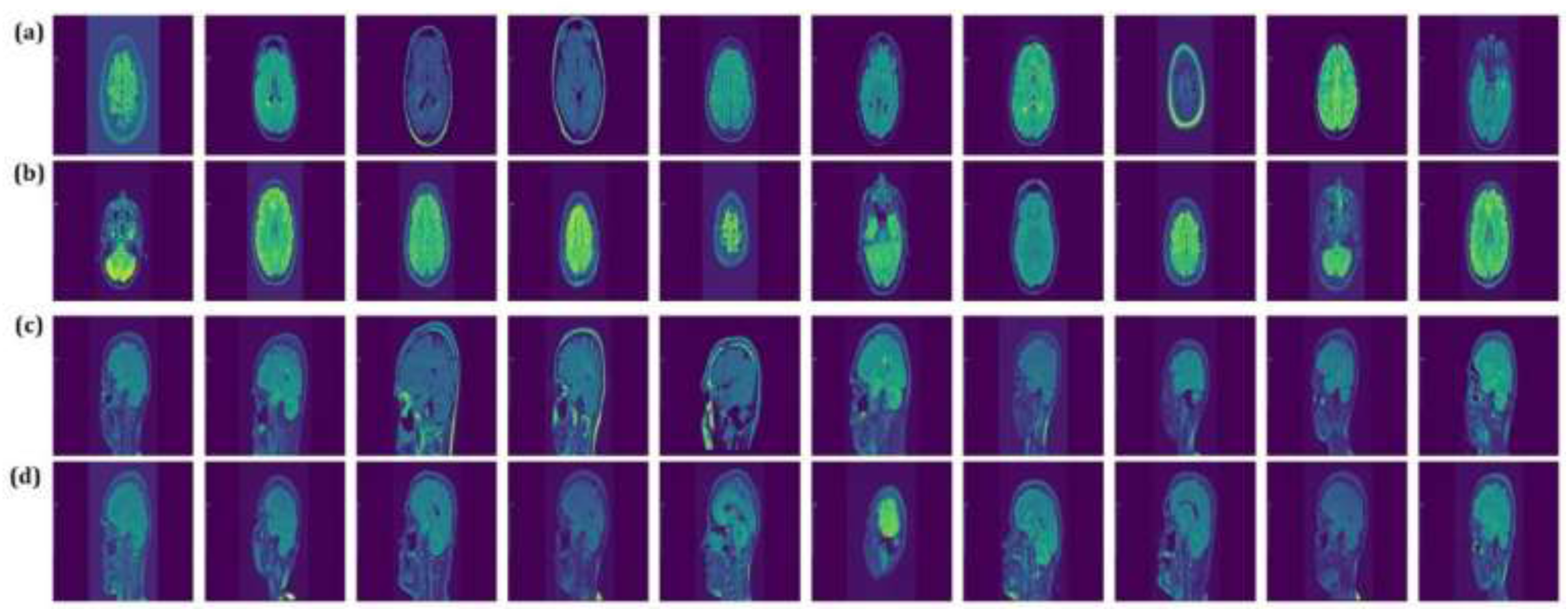
Representative MRI samples from MS and healthy groups. Randomly selected axial images from (a) MS and (b) healthy groups; Randomly selected sagittal images from (c) MS and (d) healthy groups.

We divided the dataset into training, validation, and test subsets to ensure robust model evaluation. Initially, the entire dataset was split into a combined training-validation set and a separate test set, with 20% of the data reserved for testing. This division employed stratified sampling to preserve the original class distribution across the subsets, thereby preventing class imbalance issues (38). Subsequently, the combined training-validation set was further split into distinct training and validation subsets, allocating 20% of the data for validation. Both splitting steps were conducted using a fixed random seed to ensure reproducibility of the data partitions.

For training data, augmentation was performed using Kornia, a PyTorch-based computer vision library (39). Fig. 2a and Fig. 2b depict the size of the training image data before augmentation for axial and sagittal images, respectively, while Fig. 2c illustrates the size of the training image data following augmentation.

**Fig. 2.**
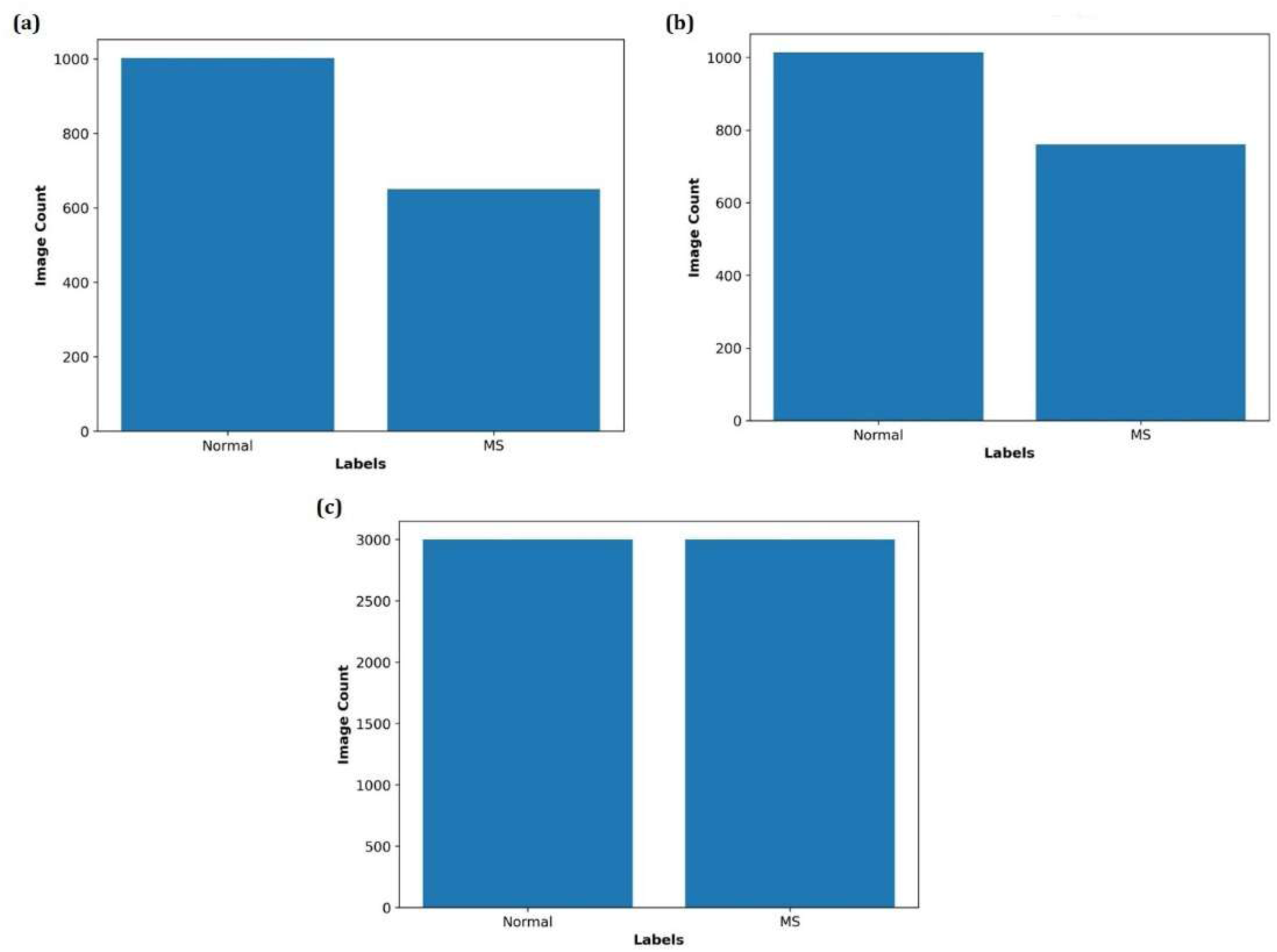
Training image dataset sizes before and after augmentation. (a) Number of axial training images before augmentation. (b) Number of sagittal training images before augmentation. (c) Number of training images after augmentation.

To standardize input dimensions for the mode, all images were resized to 224 × 224 pixels. We normalized pixel intensities to the [0.1] range by dividing by the maximum 8-bit value (255), ensuring consistent input scaling throughout the dataset.

### 3.2 Classical deep learning algorithms

#### 3.2.1 Convolutional neural network

Among DL models. convolutional neural networks are particularly prominent and widely applied in tasks such as object detection, speech recognition, image classification, and biomarker detection (40, 41). CNNs employ multi-layered architectures to learn hierarchical data representations, where each layer extracts increasingly complex features (42). This structure integrates feature extraction and classification into a single efficient framework that requires minimal preprocessing. As a result, CNNs can automatically learn and identify patterns directly from raw data without manual intervention. The core components of CNNs include convolutional layers, activation functions, pooling layers, and fully connected layers (41).

##### Convolutional layers

These layers extract distinctive features from input data through multiple convolution operations using learnable kernels (43). This process preserves spatial structure while reducing parameters through local connectivity and parameter sharing, enabling hierarchical feature learning. The output of a convolutional layer is a set of feature maps that highlight specific patterns in the data.

##### Activation functions

These functions are applied directly to the output of convolutional layers to introduce non-linearity into the model (44). This non-linearity allows CNNs to capture and represent complex, non-linear relationships within data. It enables the network to solve tasks beyond simple linear classification or regression and to effectively model intricate real-world patterns. Common activation functions include the Rectified Linear Unit (ReLU), sigmoid, and hyperbolic tangent (tanh).

##### Pooling layers

These layers usually follow convolutional and activation layers and reduce the number of network connections by downsampling and reducing the dimensionality of the input data (45). This reduction in data size lowers computational demands and aids in minimizing overfitting.

##### Fully connected (FC) layer

The FC layer is typically positioned at the end of a CNN to connect every neuron to all neurons in the preceding layer (46). It receives input from the last convolutional or pooling layer and acts as a classifier to enable the network to make predictions.

In this study, we implement a classical CNN architecture composed of a fixed convolutional base followed by a configurable dense classifier. The convolutional base includes two blocks: each consists of a Conv2d layer with ReLU activation and max pooling. Specifically, the first block applies 32 filters of size 3×3 with unit padding to the input, followed by ReLU and 2×2 max pooling with stride 2. The second block increases the number of filters to 64 while maintaining the same kernel and padding configuration. After feature extraction, spatial dimensions are flattened into a vector whose size is determined dynamically during initialization via a dummy forward pass.

The classifier module is then constructed based on this inferred input size and consists of a variable number of fully connected layers, each followed by ReLU activation and dropout regularization. The final dense layer outputs logits for two classes. Formally, given an input tensor *x*∈ℝ^3×B×*H*×*W*^, the network computes:

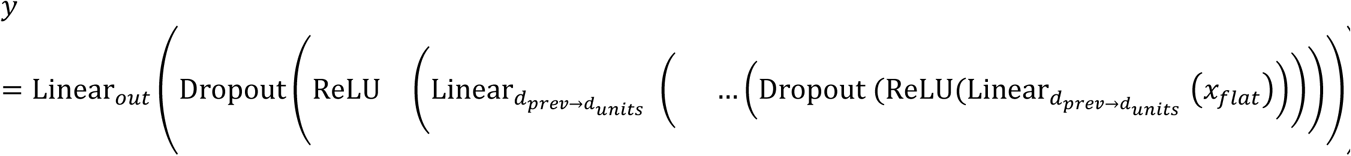

where *x*_*flat*_ is the resulting flattened feature vector with shape *B* × *d*_*in*_. *B* represents the batch size (number of input samples processed together), *d*_*in*_ denotes the flattened feature dimension output by the convolutional base. *d*_prev_ and *d*_units_ represent the input and output dimensions of each linear transformation, respectively.

This modular design supports flexible experimentation with network depth and regularization strength while maintaining a simple and interpretable baseline CNN structure.

#### 3.2.2 DenseNet121 Architecture

DenseNet (Densely Connected Convolutional Network) is a convolutional neural network architecture designed to improve feature reuse and gradient flow by establishing direct connections between all layers within a dense block (47). DenseNet121, a 121-layer variant, achieves high parameter efficiency and strong performance in image classification tasks, making it well-suitable for medical imaging applications (48). Unlike traditional CNNs, where layers receive input only from the immediately preceding layer, DenseNet121 connects each layer to every preceding layers in the same dense block by concatenating their feature maps. Formally, the output of layer ℓ is defined as:

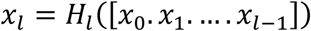

Where, [*x*_0_. *x*_1_. …. *x*_*l*−1_] denotes the concatenation of feature maps from all previous layers, and *H*_ℓ_is a composite function comprising batch normalization (*BN*), *ReLU* activation, and a 3×3 convolution (Conv_3×3_). To improve computational efficiency, a 1×1 convolution (bottleneck layer. Conv_1×1_) precedes *H*_*l*_ to reduce channel dimensionality and to improve computational efficiency:

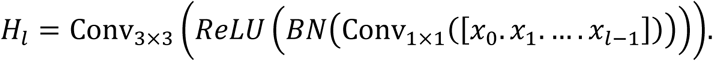

Each layer in a dense block produces *k* feature maps, where *k* is the growth rate (commonly 32), resulting an output channel count after *l* layers:

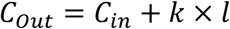

In our implementation, we used the pretrained DenseNet121 backbone from torchvision, modified to freeze the initial stem layer and the first dense block to prevent overfitting on small datasets. The final classifier layer is replaced with a configurable stack of FC layers, including ReLU activations and dropout regularization. Specifically:

- The output of the DenseNet121 backbone is passed through an adaptive average pooling layer, reducing spatial dimensions to 1×1.
- The resulting tensor is flattened into a 1024-dimensional feature vector.
- A variable number of FC layers follow, each with configurable width and dropout rate.
- The final linear layer outputs logits for two classes.

#### 3.2.3 ResNet50 Architecture

Residual Networks (ResNet) address the vanishing gradient problem by introducing residual connections that facilitate the training of deeper convolutional neural networks (49). These skip connections allow gradients to flow more directly through the network, simplifying optimization and improving learning stability. While residual connections help mitigate vanishing gradients, batch normalization and ReLU activation also play critical roles in ensuring stable and efficient training.

ResNet50, a 50-layer variant, achieves strong performance in image classification and is widely adopted in various applications due to its balance between accuracy and computational efficiency (50). The architecture begins with a stem comprising a 7×7 convolution, batch normalization, ReLU activation, and a 3×3 max-pooling layer, which transforms input images into feature maps of size 64 channels. This is followed by four stages of residual bottleneck blocks. Each block expands, transforms, and reduces channel dimensions using three convolutional layers: two 1×1 convolutions and one 3×3 convolution:

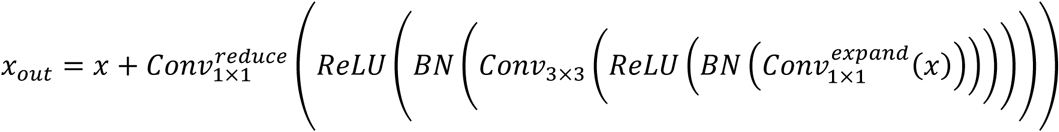

Where, Conv_1×1_^expand^ expands channels, *Conv*_3×3_ applies a 3×3 convolution, and *Conv*_1×1_^*reduce*^ reduces channels to the output dimension *C*_*out*_. A skip connection adds the input *x to the output.* with a 1×1 convolution applied to *x* if channel dimensions differ. For an input of size 224×224, the final stage of four outputs feature maps of size 7×7×2048, which are spatially reduced via an adaptive average pooling layer to a 2048-dimensional vector:

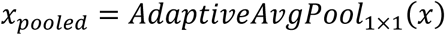

The classifier consists of a configurable stack of fully connected layers, each with ReLU activation and dropout, culminating in a final linear layer that produces class logits:

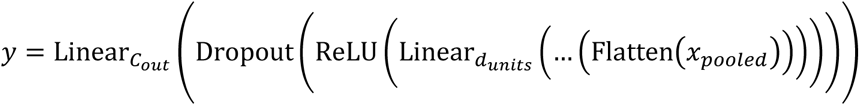

Where, Flatten converts the pooled features into a vector of size 2048 (51). To leverage transfer learning, we initialize the ResNet50 backbone using pretrained weights from a specified path. The stem and the first residual stage— including the initial convolution, batch normalization, activation, max pooling, and first residual block — were frozen to preserve low-level features learned on large-scale datasets. Later layers and the newly defined dense classifier were fine-tuned during training.

#### 3.2.4 EfficientNet-B3 Architecture

EfficientNet is a family of convolutional neural networks that optimize accuracy and efficiency by uniformly scaling network depth, width, and input resolution using a compound scaling factor (52, 53). EfficientNet-B3, a mid-sized variant, balances computational cost and performance, making it suitable for image classification tasks. The architecture is composed of a stem layer followed by a sequence of mobile inverted bottleneck convolution (MBConv) blocks augmented with squeeze-and-excitation (SE) modules (54). Each MBConv block processes input feature maps *x*∈ℝ*^H^*^×*W*×*C*^in through the following operations:

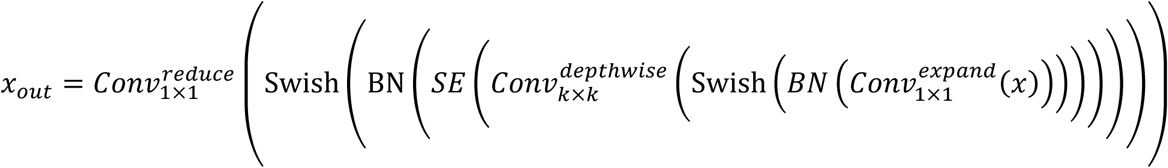

Here, *Conv*_1×1_^*expand*^ increases channel dimensionality, *Conv*_*k*×*k*_^*depthwise*^ applies a depthwise convolution, and *Conv*^*reduce*^ reduces channels. A residual skip connection adds the input *x* to the output when their dimensions’ match. The *SE* feature maps via global average pooling and two fully connected layers:

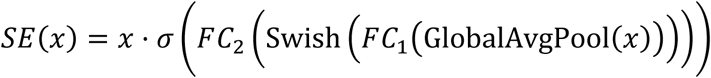

Where, GlobalAvgPool reduces spatial dimensions. *FC*_1_ and *FC*_2_ are fully connected layers and *σ* is the sigmoid function.

The EfficientNet-B3 backbone outputs feature maps of size 1536 channels for standard 224×224 RGB inputs. These feature maps are spatially reduced via global average pooling, resulting in a 1536-dimensional feature vector. The final classifier includes a configurable stack of FC layers with ReLU activation and dropout regularization, culminating in a linear output layer producing class logits:

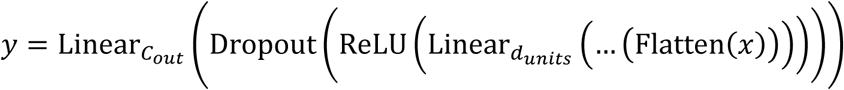

Where, flatten reshapes pooled features into a vector of size 1536.

To leverage transfer learning, we initialized the EfficientNet-B3 backbone using pretrained weights. Early layers—including the stem and initial MBConv blocks (up to block 4)—were frozen to retain pretrained features and reduced overfitting on small datasets. Later layers and the newly defined dense classifier were fine-tuned during training.

### 3.3 Hybrid classical-quantum model

In this study, we introduced hybrid classical-quantum models including QCNN, QDenseNet121, QResNet50, and QEfficientNet-B3 that integrate classical convolutional feature extraction with a variational quantum circuit to construct a quantum-enhanced classifier. The architecture is composed of three main components: (i) a classical convolutional base for spatial feature learning, (ii) a dimensionality reduction layer that maps classical features into a quantum-compatible latent space, and (iii) a parametrized quantum circuit followed by a flexible dense classifier. Fig. 3 shows the schematic flowchart of hybrid classical-quantum models architectures that are explained in further sections. Overall details of each architecture are also shown in Fig. 4.

**Fig. 3.**
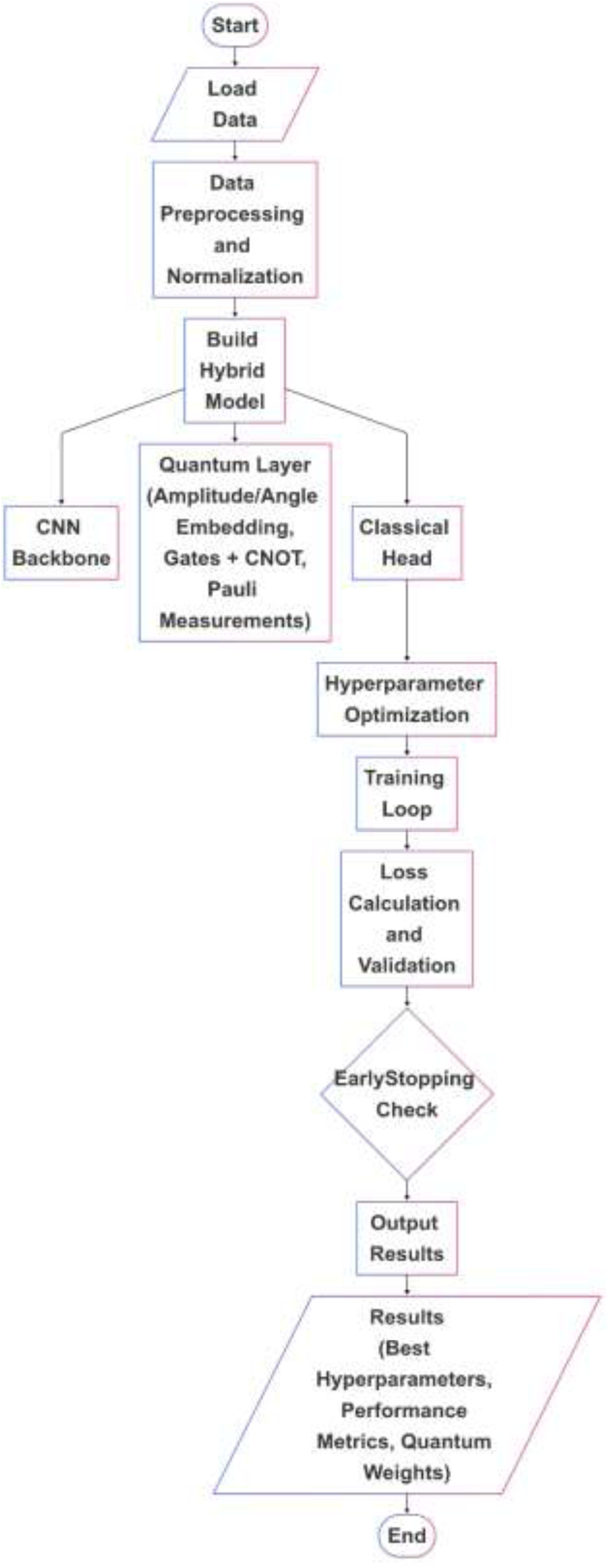
Schematic representation of the hybrid classical-quantum architectures.

**Fig. 4.**
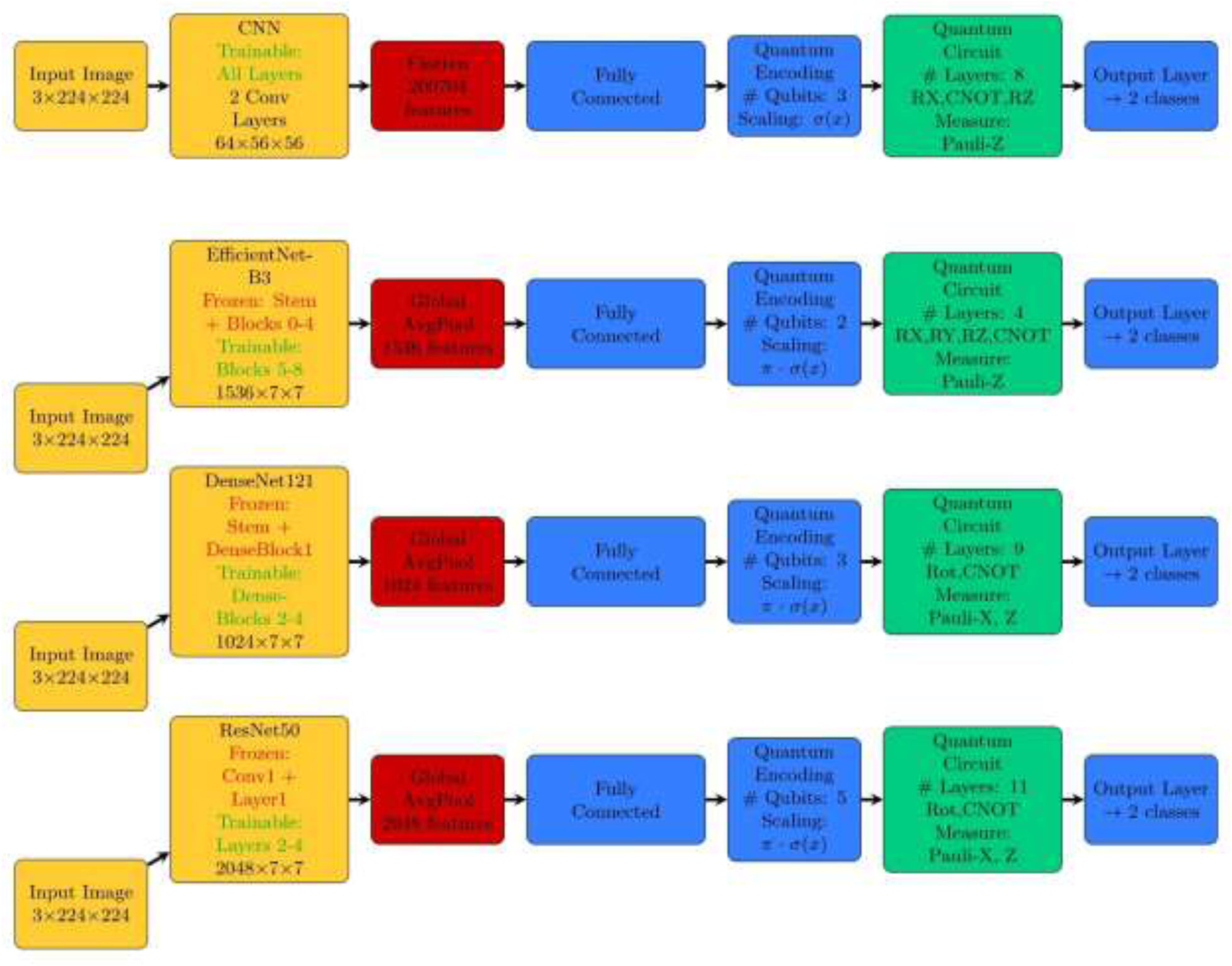
Overall details of hybrid classic-quantum architecture.

#### 3.3.1 QCNN Architecture

The QCNN model begins with a standard convolutional neural network comprising two blocks of Conv2d-ReLU-MaxPool operations. Each convolutional layer preserves spatial resolution through unit padding while increasing channel depth, followed by 2×2 max pooling that reduces spatial dimensions. This feature extraction stage ensures that the model captures hierarchical spatial patterns before transitioning to quantum processing. The output is then flattened and projected via a linear layer into a latent vector of size *n*_qubits_, matching the number of qubits in the subsequent quantum layer.

The quantum component is implemented as a parameterized quantum circuit executed on a simulated device using PennyLane (55). Each qubit undergoes an RX rotation based on the corresponding classical input, followed by multiple layers of entangling CNOT gates and trainable Z-axis rotations. Expectation values of Pauli-Z observables across all qubits are returned as the final measurement and fed into a configurable stack of dense layers. These dense layers consist of fully connected layers, each followed by ReLU activation and dropout regularization, except the final output layer which maps to 2 class logits.

The model processes input images through the following stages:

1. *Convolutional Feature Extraction:*

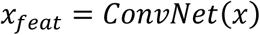

Where, *ConvNet* consists of two Conv2d-ReLU-MaxPool blocks, reducing spatial dimensions.
2. *Flattening and Dimensionality Reduction:*

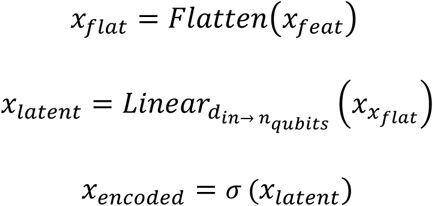

Where, *σ* denotes the sigmoid function, ensuring inputs fall within the valid range for quantum encoding and *x*_*talent*_ is obtained through a linear transformation projects the high-dimensional flattened features into a lower-dimensional space with size matching the number of qubits in the quantum circuit
3. *Quantum Circuit Layer:*

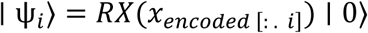

The encoded vector *x*_*encoded*_ is fed into a variational quantum circuit. Each batch element is processed independently on a simulated quantum device with n_*q*_ qubits. The circuit performs input encoding: followed by entangling CNOT gates and RZ rotations across layers. Final measurements are taken as:

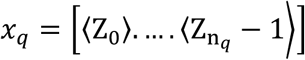
4. *Final Classification:*

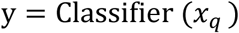

The classifier consists of a customizable sequence of fully connected layers, each followed by ReLU activation and dropout regularization, ending with a final linear layer that outputs the class logits.

#### 3.3.2 QDensenet121/QResNet50 Architecture

##### 1. Classical Feature Extraction

The backbone of QDenseNet121 or QResNet50 extract high-level feature vectors z ∈ ℝ ^1024^ and z∈ ℝ ^2048^, respectively, via adaptive average pooling of convolutional feature maps. These vectors are further processed by *n*_dense_ fully connected layers with ReLU activations and dropout regularization:

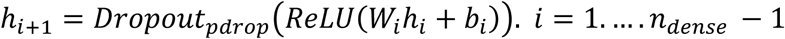

where ℎ_0_ = *z*. *W*_*i*_ ∈ ℝ^*udense*^ ^× *uin*^. *b*_*i*_ ∈ ℝ^*udense*^. *u*_*in*_ is the input dimension for the first dense layer, and *u*_*dense*_ is the number of units in each classical dense layer, thereafter. The final dense layer projects to a vector of size 2^*nqubits*^:

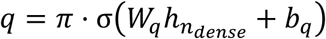

Where, σ is the sigmoid function, ensuring the quantum circuit input parameters lie within [0, π], *W*_*q*_ and *b*_*q*_ are weight matrix and bias vector projecting the final dense layer output to the quantum circuit input size 2^*nqubits*^.

##### 2. Quantum Circuit Encoding and Variational Layers

The vector q is normalized and encoded into a quantum state via amplitude embedding on an *n*_*qubits*_-qubit system:

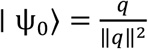

Here, ∣ ψ_0_⟩ ∈ ℂ^2^ is a normalized quantum state.

The variational quantum circuit consists of *n*_*layers*_ layers, each applying parameterized single-qubit *U*_*Rot*_(θ_*k.j*.0_. θ_*k.j*.1_. θ_*k.j*.2_) = *R*_*z*_(θ_*k.j*.2_). *R*_z_(θ_*k.j*.1_). *R*_*z*_(θ_*k.j*.0_) rotations on qubit *j*, followed by entangling CNOT gates between adjacent qubits:

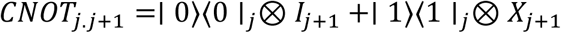

in which, *I* denotes identiy matrix and *X* indicates X rotation. The output is a vector of expectation values:

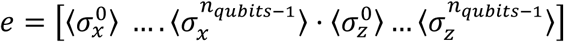

Where, σ_x_^j^ and σ_z_^j^ are Pauli operators.

##### 3. Classical Post-Processing and Classification

The quantum output e is fed into a classical classifier comprising two fully connected layers with ReLU activation and dropout:

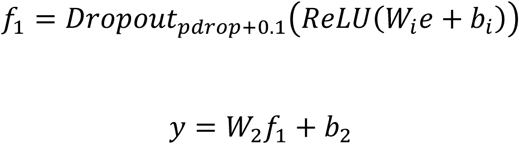

Where *W*_*i*_ ∈ ℝ^*udense* × *uin*^, *b*_*i*_ ∈ ℝ^*udense*^, *W*_2_ ∈ ℝ^2×*uclassical*^, y ∈ ℝ^2^ represents binary class logits.

#### 3.3.3 QEfficientNetB3-quantum architecture

##### 1. Classical Feature Processing

The EfficientNet-B3 backbone extracts a feature vector z∈ℝ^1536^ via global average pooling of its final convolutional output. This vector is processed through fully connected layers with ReLU activation and dropout regularization:

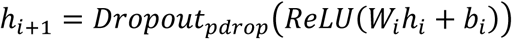

for *i* = 1. …. *n*_*dense*_ − 1, ℎ_0_ = *z*, *W*_*i*_ ∈ ℝ^*udense*^ ^× *uin*^, *b*_*i*_ ∈ ℝ^*udense*^, and *u*_*in*_ = 1536 for the first layer and *u*_*dense*_ (subsequent layers). The final layer projects to *n*_*qubits*_ dimensions (one feature per qubit) and scales to [0,π] via a sigmoid:

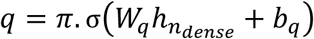

ensuring rotation angles ∈ [0. π] for quantum encoding.

##### 2. Quantum Circuit Design

The circuit employs angle embedding to map classical features to qubit rotations, reducing computational overhead compared to amplitude embedding:

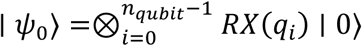

##### 3. Variational Quantum Layers

The circuit applies *n*_layers_ layers of trainable rotations and entangling gates:

Rotational Gates: Each layer *k* applies *R_x_.R_Y_.R_Z_* rotations parameterized by weights *θ_k_*_.*j.m*_:

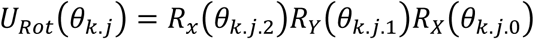

Where, *j* indexes qubits and *m* indexes rotation axes.

Entanglement: CNOT gates create a ring topology (qubit *j* entangled with (*j*+1) mod *n*_qubits_):

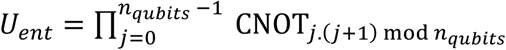

##### 4. Measurement and Classical Classifier

The circuit measures Pauli-Z expectations on all qubits:

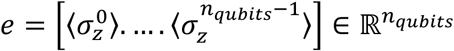

A classical fully connected network processes *e* to produce logits:

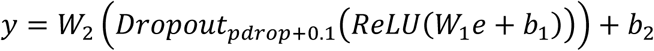

Where, W2∈R^2×dclassical^.

#### 3.3.4 Optimization

The hybrid quantum-classical models were trained using a combination of cross-entropy loss and L2 regularization to prevent overfitting and improve generalization:

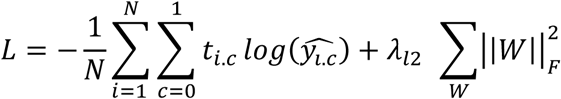

Where *t*_*i*_ ∈ {0.1}^2^ is the true label, y_*i*_ = softmax(z_*i*_) is the predicted probability, and ||*W*||^2^ is the Frobenius norm of weight matrices, using learning rate η.

To further enhance training efficiency and avoid overfitting, we implemented an early stopping mechanism that monitors validation loss during training. This ensures optimal use of computational resources while retaining the most effective model state observed throughout training. Hyperparameter tuning was carried out using Optuna, a powerful framework for automated hyperparameter optimization (56). Over 50 trials were executed across a predefined search space encompassing both classical and quantum parameters. For each trial, Optuna evaluated model performance based on validation metrics, iteratively refining its search strategy to identify the most promising configurations. The final optimal set of hyperparameters was selected based on overall performance across training, validation, and test sets. All experiments were conducted on the Leonardo supercomputer, part of the European high-performance computing infrastructure hosted by the CINECA interuniversity consortium (57). We utilized the “boost_usr_prod” SLURM partition, which provides access to NVIDIA A100 GPUs through the Leonardo Booster environment for accelerated deep learning computations. Additionally, a subset of algorithms was tested using an NVIDIA A100 GPU from a paid Google Colab Pro account to ensure development flexibility and reproducibility across environments.

## 4 Results

The performance metrics highlight the potential of hybrid classical-quantum models for enhancing MS detection, particularly in capturing complex lesion patterns. The optimized hyperparameters for each method are detailed in Table 1. Improvements in AUC, a critical metric for distinguishing MS lesions from normal tissue, were observed in QDenseNet121 and QResNet50 for axial MRI, suggesting quantum layers can enhance discriminative power in certain architectures. However, inconsistent benefits across models indicate that quantum integration is context-specific, influenced by classical model complexity and quantum circuit design. Performance declines in QCNN and QEfficientNetB3 further reveal the need for careful architectural design to maximize quantum contributions.

**Table 1.**
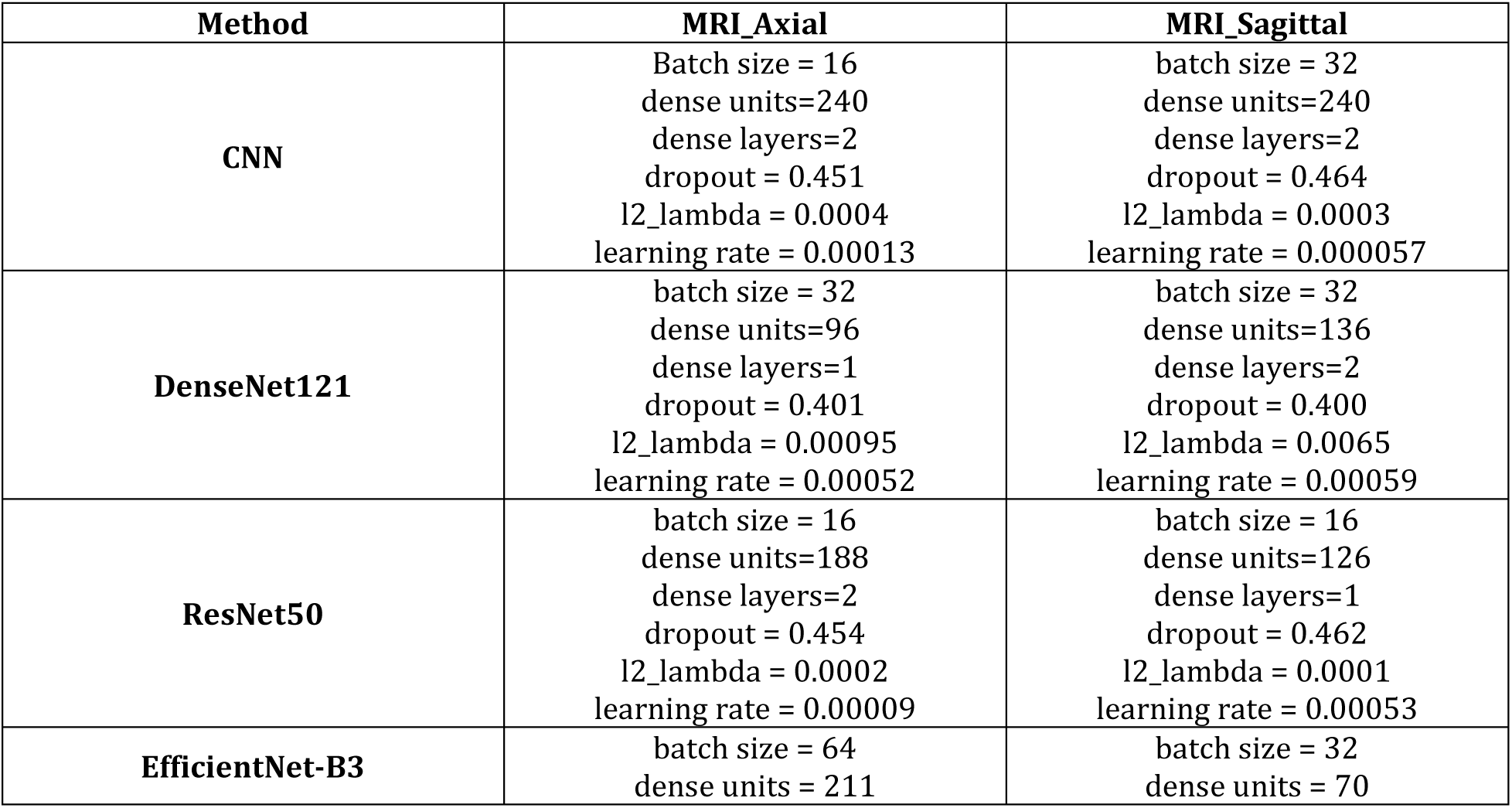
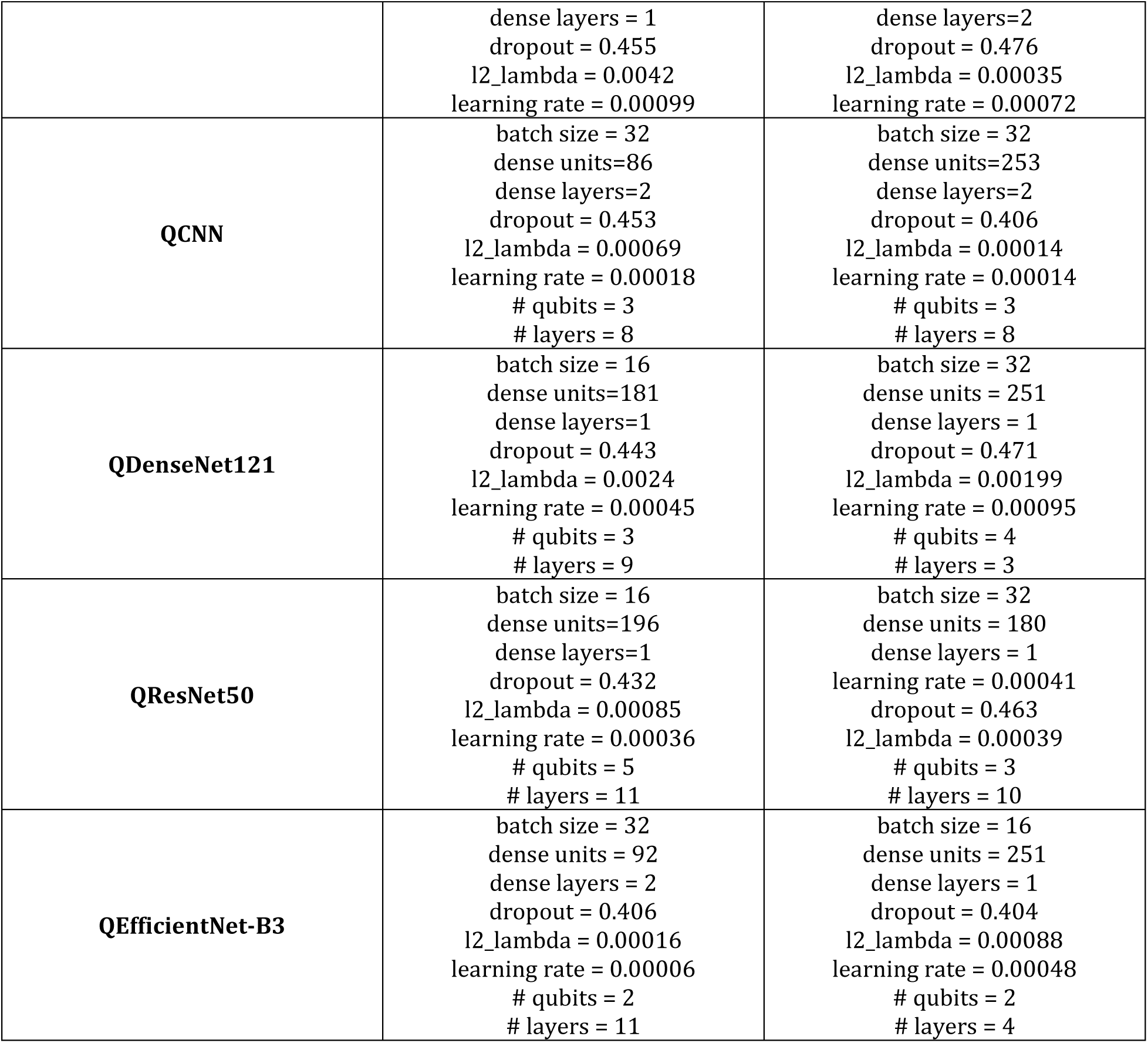
The optimized hyperparameters for each method.

| Method | MRI_Axial | MRI_Sagittal |
| --- | --- | --- |
| CNN | Batch size = 16<br>dense units=240<br>dense layers=2<br>dropout = 0.451<br>l2_lambda = 0.0004<br>learning rate = 0.00013 | batch size = 32<br>dense units=240<br>dense layers=2<br>dropout = 0.464<br>l2_lambda = 0.0003<br>learning rate = 0.000057 |
| DenseNet121 | batch size = 32<br>dense units=96<br>dense layers=1<br>dropout = 0.401<br>l2_lambda = 0.00095<br>learning rate = 0.00052 | batch size = 32<br>dense units=136<br>dense layers=2<br>dropout = 0.400<br>l2_lambda = 0.0065<br>learning rate = 0.00059 |
| ResNet50 | batch size = 16<br>dense units=188<br>dense layers=2<br>dropout = 0.454<br>l2_lambda = 0.0002<br>learning rate = 0.00009 | batch size = 16<br>dense units=126<br>dense layers=1<br>dropout = 0.462<br>l2_lambda = 0.0001<br>learning rate = 0.00053 |
| EfficientNet-B3 | batch size = 64<br>dense units = 211 | batch size = 32<br>dense units = 70 |
|  | dense layers = 1<br>dropout = 0.455<br>l2_lambda = 0.0042<br>learning rate = 0.00099 | dense layers=2<br>dropout = 0.476<br>l2_lambda = 0.00035<br>learning rate = 0.00072 |
| **QCNN** | batch size = 32<br>dense units=86<br>dense layers=2<br>dropout = 0.453<br>l2_lambda = 0.00069<br>learning rate = 0.00018<br># qubits = 3<br># layers = 8 | batch size = 32<br>dense units=253<br>dense layers=2<br>dropout = 0.406<br>l2_lambda = 0.00014<br>learning rate = 0.00014<br># qubits = 3<br># layers = 8 |
| **QDenseNet121** | batch size = 16<br>dense units=181<br>dense layers=1<br>dropout = 0.443<br>l2_lambda = 0.0024<br>learning rate = 0.00045<br># qubits = 3<br># layers = 9 | batch size = 32<br>dense units = 251<br>dense layers = 1<br>dropout = 0.471<br>l2_lambda = 0.00199<br>learning rate = 0.00095<br># qubits = 4<br># layers = 3 |
| **QResNet50** | batch size = 16<br>dense units=196<br>dense layers=1<br>dropout = 0.432<br>l2_lambda = 0.00085<br>learning rate = 0.00036<br># qubits = 5<br># layers = 11 | batch size = 32<br>dense units = 180<br>dense layers = 1<br>learning rate = 0.00041<br>dropout = 0.463<br>l2_lambda = 0.00039<br># qubits = 3<br># layers = 10 |
| **QEfficientNet-B3** | batch size = 32<br>dense units = 92<br>dense layers = 2<br>dropout = 0.406<br>l2_lambda = 0.00016<br>learning rate = 0.00006<br># qubits = 2<br># layers = 11 | batch size = 16<br>dense units = 251<br>dense layers = 1<br>dropout = 0.404<br>l2_lambda = 0.00088<br>learning rate = 0.00048<br># qubits = 2<br># layers = 4 |

### 4.1 Evaluation of Classic and Hybrid Classic-Quantum DL Models for Axial MRI

The metrics including accuracy, precision, recall, F1-score, and AUC offer insights into the models’ effectiveness in identifying MS-related features in axial MRI images (Table 2). The results reveal both the strengths of classic DL models and the potential of quantum-enhanced approaches with implications for advancing MS diagnostics.

**Table 2.**
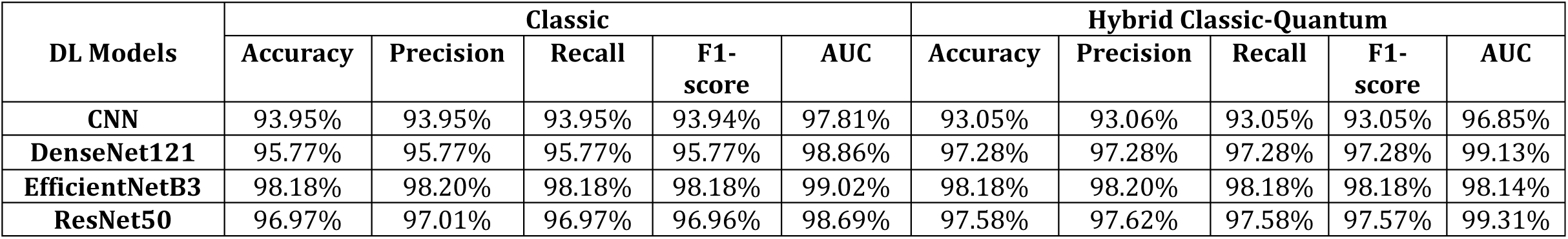
Performance metrics of classic and hybrid classic-quantum algorithms for axial MRI images.

Among the classic DL models, EfficientNetB3 achieved the highest performance, with an accuracy of 98.18%, precision of 98.20%, recall of 98.18%, F1-score of 98.18%, and an AUC of 99.02%. This performance is likely due to EfficientNetB3’s compound scaling approach, which optimizes network architecture to capture the subtle and heterogeneous patterns of MS lesions. ResNet50 and DenseNet121 also performed robustly, with accuracies of 96.97% and 95.77%, respectively, and AUC values above 98%, indicating strong discriminative ability for MS-related features. In contrast, the CNN model showed the lowest performance (accuracy of 93.95%. AUC of 97.81%), suggesting that simpler architectures may struggle to model the complex characteristics of MS lesions.

The hybrid classic-quantum models demonstrate varied performance relative to their classic counterparts. Notably, QDenseNet121 showed a significant improvement, with an accuracy increasing from 95.77% to 97.28% and AUC from 98.66% to 99.13%. Similarly, QResNet50 improved accuracy versus classical counterpart (from 96.97% to 97.58%) and AUC (from 98.69% to 99.31%), indicating that quantum circuits may enhance the model’s ability to capture residual features critical for identifying MS-specific patterns. However, the QCNN model exhibited a slight performance decline (accuracy drops from 93.95% to 93.05%. AUC from 97.81% to 96.85%), possibly due to the limited capacity of the CNN architecture to leverage quantum enhancements effectively. Intriguingly, EfficientNetB3’s hybrid model shows no improvement in accuracy (98.18%) and a slight AUC decrease (from 99.02% to 98.14%), suggesting that its highly optimized classic architecture may already be near the performance, leaving little room for quantum contributions.

### 4.2 Evaluation of Classic and Hybrid Classic-Quantum DL Models for Sagittal MRI

For sagittal MRI images, the classic DL models demonstrated exceptional performance, with ResNet50 achieving the highest metrics accuracy, precision, recall, and F1-score all at 99.15%, and an AUC of 99.93% (Table 3). This performance likely reflects ResNet50’s ability to capture deep residual features, which are critical for identifying MS lesions in the sagittal plane, where lesions may appear as irregularly shaped hyperintensities, particularly in the spinal cord or periventricular regions.

**Table 3.**
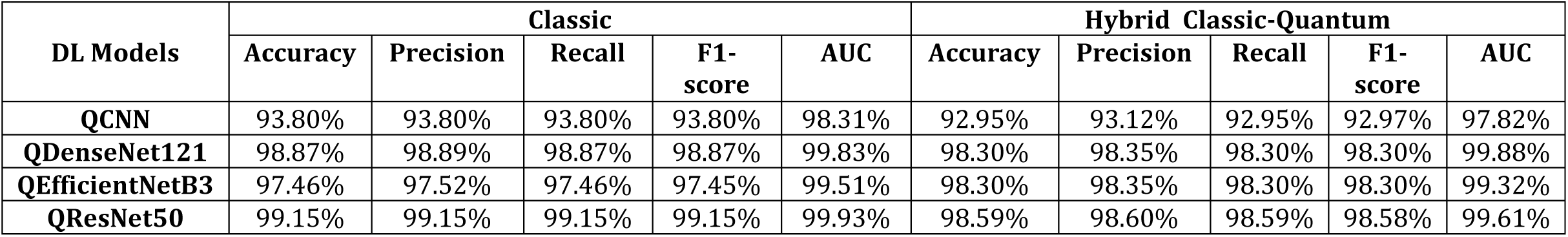
Performance metrics of classic and hybrid classic-quantum algorithms for sagittal MRI images.

DenseNet121 has the next position, with an accuracy of 98.87%, precision of 98.89%, recall of 98.87%, F1-score of 98.87%, and an AUC of 99.83%, demonstrating its strength in leveraging dense connectivity to model complex lesion patterns. EfficientNetB3, while still highly effective, showed slightly lower performance (accuracy of 97.46% and AUC of 99.51%), possibly due to its scaling strategy being less optimized for the specific spatial characteristics of sagittal images. The CNN model lags behind, with an accuracy of 93.80% and an AUC of 98.31%, indicating that simpler architectures struggle to capture the nuanced features of MS lesions in this plane.

The hybrid classic-quantum models showed mixed results compared to their classic counterparts. QDenseNet121 model slightly underperformed, with accuracy decreasing from 98.87% to 98.30% and AUC slightly increasing from 99.83% to 99.88%. This suggests that quantum enhancements may not significantly improve DenseNet121’s already strong performance. However, the AUC of QDenseNet121 (99.83%) was slightly better than classical counterpart (99.88%), indicating potential for better discriminative power. QEfficientNetB3 model showed a notable improvement in accuracy (from 97.46% to 98.30%) but a slight AUC decrease (from 99.51% to 99.32%), suggesting that quantum layers enhance overall classification, but may slightly reduce the model’s ability to rank positive cases. QResNet50 exhibited a performance decrease across all metrics (accuracy from 99.15% to 98.59% and AUC from 99.93% to 99.61%), indicating that quantum integration may disrupt the model’s finely tuned residual learning for sagittal images. The QCNN model also performed worse (accuracy drops from 93.80% to 92.95% and AUC from 98.31% to 97.82%), reinforcing that simpler architectures may not benefit from quantum enhancements, possibly due to limited quantum circuit complexity.

## 5 Discussion

In this study, we investigated the detection of multiple sclerosis lesions in axial and sagittal MRI scans using classical and hybrid quantum-classical deep learning models. Given the diverse spatial and textural patterns of MS lesions, the adaptability of these models to such variations is critical for accurate diagnosis. Our comparative analysis reveals distinct performance trends across architectures and imaging planes, highlighting the potential and limitations of quantum-enhanced DL in medical imaging.

Recent advances in deep learning have significantly improved the classification of MS using MRI data, with numerous studies reporting high-performance metrics (Table 4).

**Table 4.** Comparative overview of previous studies.

| Method | Dataset | Performance Metrics (%) | Ref |
| --- | --- | --- | --- |
| ExMPLPQ | **MS:**<br>1411 MRI from 72 patients<br>**Healthy:**<br>2016 MRI from 59 subjects | **Axial**<br>Accuracy: 98.37<br>Sensitivity: 96.46 | Macin et al. (37) |
|  |  | Specificity: 99.60<br>**Sagittal**<br>Accuracy: 97.75<br>Sensitivity: 95.01<br>Specificity: 99.80 |  |
| MSNet | **MS**<br>706 MRI from 128 patients<br>**Myelitis**<br>667 MRI from 131 patients<br>**Healthy**<br>1373 MRI from 150 subjects | Accuracy: 97.13<br>Precision: 97.23<br>Recall: 97.22<br>F1-score: 97.23 | Tatli et al.<br>(58) |
| Six-layer CNN alongside stochastic pooling. | **MS**<br>676 MRI from 28 patients<br>**Healthy**<br>681 MRI from 26 subjects | Accuracy: 95.82<br>Sensitivity: 95.98<br>Specificity: 95.67<br>Precision: 95.66<br>F1-Score: 95.81 | Wang et al.<br>(20) |
| CNN featuring wavelet pooling. | **MS**<br>676 MRI from 38 patients<br>**Healthy**<br>615 MRI from 20 subjects | Accuracy: 98.92<br>Sensitivity: 99.20<br>Specificity: 98.33<br>Precision: 99.20 | Alijamaat et al. (21) |
| D-CNN model inception V3 | **DS1:**<br>**MS**<br>1411 MRI from 72 patients<br>**Healthy**<br>2016 MRI from 59 subjects<br>**DS2:**<br>**MS**<br>1263 MRI from 60 patients<br>**Healthy**<br>2016 MRI from 59 subjects<br>**DS3:**<br>**MS**<br>1581 MRI from 20 patients<br>**Healthy**<br>2016 MRI from 59 subjects | mAP (mean Average Precision) values: 86.20, 93.77, 94.18, and 90.46 for DS1. DS2. DS3. and DS4 | Haggag et al.<br>(59) |
|  | **DS4:**<br>**MS**<br>1805 MRI from 38 patients<br>**Healthy**<br>2016 MRI from 59 subjects |  |  |
| Exemplar<br>MobileNetv2.<br>IMrMr. kNN | **MS**<br>1411 MRI from 144 patients<br>**Healthy**<br>2016 MRI from 106 subjects | **Axial:**<br>Accuracy: 99.76<br>**Sagittal:**<br>Accuracy: 99.48 | Ekmekyapar<br>and Tascı<br>(60) |

Macin et al. achieved an accuracy exceeding 98.37% for axial and 97.75% for sagittal MRI images using ExMPLPQ, a method that integrates exemplar-based multi-parametric Local Phase Quantization (LPQ) features with a k-nearest neighbors (kNN) classifier (37). This method affirmed the enduring effectiveness of handcrafted feature extraction in MS diagnosis. In another study, Tatli et al. developed MSNet, a hybrid MS classification model that innovatively merges deep learning (DenseNet201 and ResNet50) with feature engineering (NCA, ReliefF, and Chi2) and ensemble learning (SVM/kNN + majority voting) (58). By extracting and refining deep features through transfer learning and iterative voting, the model achieved 97.13% accuracy, demonstrating the advantages of combining deep neural networks with classical machine learning techniques for medical image analysis. Further improvements were observed in studies utilizing CNNs with innovative pooling strategies, such as Wang et al. (20) and Alijamaat et al. (21), who reported accuracies of 95.82% and 98.92%, respectively, reinforcing the effectiveness of convolutional architectures in MS classification. Haggag et al. employed a D-CNN model based on Inception V3 across four datasets containing MRI scans from MS patients and healthy controls, achieving mean average precision (mAP) values of 86.20%, 93.77%, 94.18%, and 90.46% for various datasets (59). Meanwhile. Ekmekyapar and Tascı utilized a hybrid approach combining Exemplar MobileNetv2 with IMrMr feature selection and kNN, attaining accuracy 99.76% on axial images and 99.48% on sagittal images (60).

In our study, EfficientNetB3 achieved the highest performance among classical models for axial MRI images, with an accuracy of 98.18% and an AUC of 99.02%. This considerable performance is attributed to EfficientNetB3’s compound scaling approach, which systematically optimizes network depth, width, and resolution to capture subtle and heterogeneous MS lesion patterns (61). ResNet50 and DenseNet121 also demonstrated robust performance, with accuracies of 96.97% and 95.77%, respectively, and AUC values exceeding 98%. Their success can be attributed to residual learning and dense feature reuse, which enhance hierarchical feature extraction (49, 53). In contrast, the simpler CNN architecture shows the lowest performance, reflecting its limited capacity to model the complex lesion patterns.

For sagittal MRI, ResNet50 excelled with near-perfect metrics (accuracy: 99.15% and AUC: 99.93%). DenseNet121 followed closely with an accuracy of 98.87% and an AUC of 99.83%, leveraging dense connectivity to integrate multi-scale features. EfficientNetB3, while still highly effective, showed slightly lower performance (accuracy 97.46% and AUC 99.51%), suggesting its scaling strategy may be less optimal for sagittal-specific features. The CNN again underperformed, reinforcing the need for deeper architectures in MS lesion detection.

The hybrid classic-quantum models introduce additional complexity to traditional deep learning architectures, resulting in varied performance across MRI planes and models, dependent on the classical backbone and imaging plane.

For axial MRI, QDenseNet121 improved accuracy by 1.51% (95.77% → 97.28%) and AUC by 0.47% (98.66% → 99.13%), likely due to quantum-enhanced feature disentanglement via amplitude embedding. The variational quantum circuit’s non-linear transformations may refine high-dimensional features, improving lesion separability. QResNet50 showed a 0.61% accuracy gain (96.97% → 97.58%) and a 0.62% AUC improvement (98.69% → 99.31%), suggesting quantum entanglement enhances discriminative power for residual features. QEfficientNetB3 exhibited no accuracy improvement (98.18%) and a slight AUC drop (99.02% → 98.14%), indicating its classical optimization may saturate performance. The QCNN model experienced a modest performance decline, with accuracy dropping from 93.95% to 93.05% and AUC decreasing from 97.81% to 96.85%, suggesting that simpler classical backbones may lack the representational capacity to fully leverage quantum augmentation.

For Sagittal MRI, QEfficientNetB3 improved accuracy by 0.84% (97.46% → 98.30%) but slightly reduced AUC (99.51% → 99.32%), suggesting a trade-off between classification and ranking performance. QResNet50 and QDenseNet121 showed modest, possibly due to interference with pre-trained residual/dense features critical for sagittal lesions. QCNN model also underperformed in sagittal MRI, reinforcing the notion that simpler architectures struggle to benefit from quantum enhancements, possibly due to limited quantum circuit complexity and classical representational power.

The hybrid models employed two primary quantum feature encoding strategies. First, amplitude embedding used in QDenseNet121 and QResNet50. maps classical feature vectors q∈ ℝ^*nqubits*^ to a normalized quantum state ∣ψ_0_⟩= q∥(q∥)_2_. Theoretical analyses show that amplitude embedding can exponentially compress high-dimensional data into quantum states; however, it requires O(2^n^) classical preprocessing, which limits scalability (62). Second, angle embedding, utilized in QEfficientNetB3, encodes n features as qubit rotations *nqubits*−1 (i)|0⟩, offering better scalability with complexity O(n), but it may lose some global correlations present in the data (63).

The quantum circuits incorporate parametrized rotation gates, which are optimized via backpropagation. These gates introduce quantum-specific non-linear transformations, potentially enhancing feature extraction, as seen in QDenseNet121’s AUC improvement for axial MRI. CNOT gates in a ring topology create entanglement between adjacent qubits, enabling quantum correlations that may capture long-range dependencies across distant image regions, relevant for MS detection due to lesions’ heterogeneous spatial distributions. This mechanism could contribute to QResNet50’s AUC gain, though QEfficientNetB3’s AUC drop suggests context-specific benefits limited by circuit design (e.g., qubit count, entanglement scope).

Measurement of the quantum states is performed via Pauli observables ⟨σ_x_⟩ and ⟨σ_z_⟩. The ⟨σ_z_⟩ observable encodes local qubit polarization, which is sensitive to lesion intensity variations, while ⟨σ_x_⟩ captures coherence between lesions and healthy tissue. The classical fully connected layers then map the measured quantum features to output predictions, with quantum noise such as shot noise being regularized through dropout techniques.

Despite these promising results, several challenges limit the clinical applicability of hybrid classical-quantum models. Current quantum hardware constraints restrict the number of qubits. impacting the dimensionality of the feature space that can be encoded. Additionally, clinical MRI data often contain artifacts. such as motion blur and partial volume effects, which can degrade quantum coherence and computational fidelity. Future work should prioritize error mitigation strategies. such as dynamical decoupling and probabilistic error cancellation, to enhance robustness under noisy conditions. Furthermore. the theoretical framework for quantum advantage in medical imaging remains underdeveloped, with rigorous performance bounds yet to be established. Future research should focus on deriving rigorous mathematical frameworks to quantify the conditions under which quantum enhancements provide measurable improvements over classical approaches. Addressing these challenges will be essential for translating quantum-based diagnostic tools from theoretical constructs into clinically viable solutions.

## Conclusion

This study demonstrates that classical deep learning architectures achieve robust performance in MS lesion detection across different MRI planes, with residual and densely connected networks showing particular effectiveness. Hybrid quantum-classical models provide improvements, enhancing certain architectures through quantum-enabled feature refinement while offering no advantage for simpler networks. The benefits of quantum integration prove highly dependent on both the base model’s design and the specific imaging plane being analyzed. While quantum circuits show promise for medical image analysis through their unique feature encoding and transformation capabilities, their current utility remains constrained by fundamental limitations in scalability and noise susceptibility. Future progress in this emerging field will depend on co-designed quantum-classical systems.

## Declarations

### Human Ethics and Consent to Participate

Not applicable.

## Competing interests

I declare that the authors have no competing interests as defined by Springer, or other interests that might be perceived to influence the results and/or discussion reported in this paper.

## Funding

No funding.

## Availability of data and materials

All data generated or analysed during this study are included in this published article [and its supplementary information files].

## Authors’ contributions

M-ZG implemented and executed all deep learning code, interpreted and visualized the results, drafted the manuscript, and supervised the project. EA investigated the project and contributed to writing the manuscript. All authors reviewed and approved the final version of the manuscript.

## Acknowledgements

I would like to acknowledge support from the ICTP through the Associates Programs (2023–2028).

## Notes

### Competing Interest Statement

The authors have declared no competing interest.

